# Non-pharmaceutical interventions for persons living with young-onset dementia and their informal caregivers: a systematic review with meta-analysis

**DOI:** 10.1101/2025.09.04.25335102

**Authors:** Magdalena Vogt, Nicole Helfenberger, Christian Appenzeller-Herzog, Laura Adlbrecht, Julian Hirt

**Affiliations:** Department of Health, Eastern Switzerland University of Applied Sciences, St. Gallen, Switzerland; University Medical Library, University of Basel, Basel, Switzerland; Pragmatic Evidence Lab, Research Center for Clinical Neuroimmunology and Neuroscience Basel (RC2NB), University Hospital Basel and University of Basel, Basel, Switzerland; Institute of Health, Midwifery and Nursing Science, Medical Faculty, Martin Luther University Halle-Wittenberg, Halle (Saale), Germany

**Keywords:** Young-onset dementia, Dementia, Frontotemporal dementia, Caregivers, Non-pharmaceutical interventions, Systematic review

## Abstract

To investigate the effects of non-pharmaceutical interventions for persons living with young-onset dementia (YOD) including frontotemporal dementia and their informal caregivers, we conducted a systematic review including randomized and non-randomized controlled trials. We searched bibliographic databases and performed citation and web searches. We found nine trials assessing interventions on education and information or support and counselling that were published between 1990 and 2024 (median sample size: 58). Meta-analyses revealed no statistically significant impact on behavioral outcomes, activities of daily living and quality of life of persons living with YOD and no statistically significant impact on burden, depression and anxiety, and quality of life of informal caregivers. Evidence on the effectiveness of non-pharmaceutical interventions for persons living with YOD and their informal caregivers is limited and inconsistent. Further, larger, and multiple randomized controlled trials assessing the impact of non-pharmaceutical interventions with comparable outcomes, standardized measurements, and longer follow-ups are needed.

**HIGHLIGHTS:**

- Young-onset dementia (YOD) including frontotemporal dementia (FTD) is challenging for individuals and families.
- For education and information interventions, there was no statistically significant impact on behavioral outcomes (SMD −0.18, 95% CI −0.67; 0.31), activities of daily living (SMD 0.05, 95% CI −0.45; 0.54) and quality of live (MD −8.93, 95% CI −30.73; 12.86) of persons living with YOD.
- For support and counselling interventions, there was no statistically significant impact on burden (SMD −0.09, 95% CI −0.66; 0.47), depression and anxiety (SMD −0.20, 95% CI −0.66; 0.25), and quality of live (SMD −0.56, 95% CI −2.12; 1.00) of informal caregivers.

## INTRODUCTION

Young-onset dementia (YOD) refers to any dementia diagnosed before the age of 65.^1^ With an estimated prevalence of 119 per 100’000, YOD accounts for about five percent of all dementia cases.^2^ Common symptoms in persons living with YOD are memory complaints, depression, behavioral issues, and physical symptoms such as gait disturbance, seizures, peripheral neuropathy, and visual impairment. Frontotemporal dementia (FTD) is a specific form of YOD that is characterized by progressive deficits in behavior, executive function, or language.^3^ Being diagnosed with YOD can lead to significant consequences for a younger person, including early retirement, financial implications, and the psychological challenge of coping with cognitive decline.^4^ Also family members of persons living with YOD experience emotional and psychological challenges and unmet needs pre and post diagnosis,^4,5^ characterized by worries about dependency, anxiety, and increased depression.^6^ Despite these challenges, current interventions do not meet the personal and psychological needs of persons living with YOD and their informal caregivers,^7^ as there exist YOD-specific barriers including ineligibility, unaffordability, lack of security and lack of childcare. Therefore, post-diagnostic care for persons living with YOD should be tailored, flexible, affordable and provide meaningful engagement.^8^ From a qualitative systematic review, Zhang et al. (2025) derived three themes of supportive care needs of persons living with YOD: empowerment through knowledge and planning, promotion of physical and psychosocial well-being, and the need for a strong support network.^9^

Various interventions aim to support activities of daily living, reduce neuropsychiatric symptoms, slow cognitive decline and improve social engagement for both persons living with YOD and their informal caregivers.^10^ There can be benefits in improving quality of life in persons living with YOD and their informal caregivers through these non-pharmaceutical interventions, e.g. in reducing care burden and psychological stress, improved self-esteem and sense of purpose.^10,11^ A systematic assessment of the benefits and harms of non-pharmacological interventions for persons living with YOD and their informal caregivers is currently lacking. The objective of this study was therefore to investigate the effects of non-pharmaceutical interventions for persons living with YOD and their informal caregivers and to explore the intervention characteristics.

## METHODS

We performed a systematic review following our pre-specified methods as registered on the international prospective register of systematic reviews (PROSPERO, CRD42025645744).^12^ We followed the “Preferred Reporting Items for Systematic reviews and Meta-Analyses” (PRISMA) 2020 statement to structure this report.

### Eligibility criteria

We included primary studies with a comparative interventional design (i.e. participants allocated to one of two or more groups, e.g., individual or cluster randomized controlled trial, non-randomized controlled trial, stepped-wedge design) in any phase (e.g. pilot, feasibility, evaluation) with the following characteristics:

- Population: Persons living with YOD, FTD, and/or their informal caregivers. YOD is typically being defined as onset of dementia until the age of 65. However, we do not consider any age-related cut-off but rely on the terminology as used by study authors (e.g., early-onset or young-onset dementia). Studies including persons living with YOD, FTD, and/or their informal caregivers as a subgroup among other types of dementia were included if they presented information on a separate analysis for our target population in the abstract.
- Intervention and comparison: Any post-diagnostic non-pharmaceutical intervention (e.g., care, nursing, medical, therapeutical, rehabilitative, supportive, behavioral, environmental, social, and system intervention) with any comparison (e.g., active, sham, usual care). Drug interventions, dietary supplements, and diagnostic interventions were not considered.
- Outcome: Any clinical outcome measured in persons living with YOD or informal caregivers. We did not consider process evaluation outcomes (e.g., satisfaction with the intervention, feasibility of the intervention, other implementation issues), biomarkers (except from digital biomarkers), and surrogate outcomes (e.g., vital signs, labor measures, imaging measures, electrocardiogram, or ultrasound).
- Setting: Any country, context, or setting.

We considered journal articles following an Introduction-Methods-Results-and-Discussion (IMRaD) structure published in English or German for inclusion with no restrictions to the publication year.

Evidence syntheses and study protocols were excluded but kept as seed reference for citation searching (see below for details).

### Information sources and search strategy

We searched the following databases on February 7, 2025: MEDLINE/Ovid, Embase/Ovid, CINAHL/EbscoHost, CENTRAL/Cochrane Library, and PsycInfo/Ovid. The database search strategy was developed by two reviewers (MV and JH) with consultation of a medical information specialist (CAH); informed by population-related search terms identified in relevant evidence syntheses (e.g., young-onset, early onset, younger onset, or frontotemporal combined with dementia or Alzheimer) combined with search filters for study design (i.e., clinical trials using Canadian Agency for Drugs and Technologies in Health (CADTH) ‘All Clinical Trials’ filters; except for CENTRAL) and publication type (i.e., full publication of study findings, only for Embase and CENTRAL) and a NOT-component (aiming at excluding biomedical and basic research). The search strategy was drafted for the search in MEDLINE and translated to the remaining databases with the support of Polyglot Search^13^ (Appendix 1). Deduplication was performed using Citavi.^14^

Using the new web application Co*Citation Network (https://CoCitationNetwork.github.io), we performed citation searches^15^ on March 13, 2025 using eligible study reports,^16–28^ pertinent evidence syntheses,^7,10,11,29^ and study protocols^20,30,31^ as retrieved by the database search as seed references. Specifically, we conducted unranked direct citation searching (UDCS; for cited and citing references) and ranked (in)direct citation searching (RICS; for top-ranking cited, citing, co-cited and co-citing references) as described in detail elsewhere.^32^ One iteration of citation searching was performed. Deduplication (against references retrieved by database searching and within but not between citation searching methods [RICS and UDCS]) was performed using a method that is largely identical to the Bramer method and has been documented in an online video tutorial (https://osf.io/jaeu5). After finishing the search and screening of references retrieved by database and citation searching, we searched Google Scholar using specific search terms (e.g., early-onset, young-onset, frontotemporal dementia, non-pharmaceutical intervention) and considered the first 20 search hits. We also made use of artificial intelligent search engines on April 29, 2025 (undermind.ai [free version] and Elicit.com [Plus version]) by prompting our review question (i.e., “What are the effects of non-pharmaceutical interventions for persons living with young-onset or frontotemporal dementia and their caregivers?”) and screening the first eight hits each.

Finally, one reviewer (MV) checked the reference lists of reviews^33–38^ that were retrieved from citation searching for additional eligible studies.

### Study selection

Two reviewers (MV and NH) independently screened titles, abstracts, and full-texts using the Rayyan web-app.^39^ Any conflicts were discussed between the two reviewers, or with consultation of a third reviewer (JH). Results retrieved by searches on Google Scholar and from artificial intelligent search engines were screened by one reviewer (MV), with consultation of a second reviewer (JH) in any unclear case.

### Data extraction

We used Elicit, an artificial intelligent (AI) research assistant,^40^ to extract data from eligible studies. All extractions were verified or complemented by a human reviewer (MV). Extracted outcome and results data were in addition verified by a second human reviewer (NH or JH). We extracted the following information: general study characteristics (corresponding author, protocol link, registry number, publication date, DOI), study design, design characteristics (number of study arms, number study centers, allocation type), study objective, population (type of participants, sample size, age and sex of participants, type of dementia), country of conduct, study setting, intervention (type and name of intervention(s) and comparator(s), duration, facilitator, frequency, content, delivery and location), outcome (name/type, participant group, measurement, measurement type and direction, timepoint(s) of measurement, maximum length of follow-op), and results (type, group differences, total number of experimental and control group, mean and SD of groups, latest follow-up for all intervention groups per outcome, narrative and numeric results of intervention effects).

### Assessment of the methodological quality

One reviewer (MV) assessed the risk of bias using the Mixed Methods Appraisal Tool (MMAT) Version 2018^41,42^ (relevant categories of study: quantitative randomized controlled trials and quantitative non-randomized) with consultation of a second reviewer (JH) in any unclear cases. Signalling questions were answered with yes, no, or can’t tell. Can’t tell meant that there was insufficient information to answer with yes or no. The appraisals were double-checked by a second reviewer (NH; Table 3). We did not assess the certainty of evidence.

### Data analysis and synthesis

To describe the study and intervention characteristics, we performed a narrative summary report based on tabulated data extractions using numbers and percentages. This was based on meaningful groupings of study characteristics, i.e., interventions, and outcomes; based on clinical and methodological expertise of three reviewers (MV, LA, and JH; Appendix 3).^43^

To provide an overview of the effects of non-pharmaceutical interventions for persons living with YOD and their informal caregivers, we designed a harvest plot showing effect directions of all study outcomes and results and we narratively summarized its content. Harvest plots provide an overview of evidence from studies with multiple designs reporting heterogenous interventions, outcomes, and other clinical characteristics with differential intervention effects.^44^ Interventions and outcomes for the harvest plot and narrative synthesis were inductively categorized by two reviewers (JH, MV).

Three authors (MV, LA, and JH) explored the clinical heterogeneity and thus the fundamental potential for statistical meta-analyses, based on discussion on the similarity and differences in study characteristics (i.e., population, intervention, comparison, and outcome). This assessment informed the meaningfulness of potential statistical meta-analyses. Statistical heterogeneity was explored based on visual results of forest plots to consider the direction and magnitude of effects and the degree of overlap between confidence intervals. In addition, we based our interpretation on the I^2^ statistics.^45^ As a result from these clinical and statistical explorations and assessments, we performed pre-specified explorative meta-analyses with outcome results for persons living with dementia (behavior outcomes, activities of daily living, and quality of life) and informal caregivers (burden, depression, and quality of life), calculated by the free web-based tool MetaAnalysisOnline.com.^46^ We used a random effects model, the inverse variance method, hedges’ g for estimating SMD, and restricted maximum likelihood for estimating between study heterogeneity. We pooled the intervention effects of the experimental group versus the control group on different outcomes with the standardized mean difference (SMD) using 95% confidence intervals (CI) based on mean, standard deviation (SD), and number of participants.^45^ Where the SD was not available, we calculated it from the reported CI using RevMan Web.^47^ For SMD calculation and to ensure that all the scales point in the same direction (i.e., lower values indicate a more positive impact) and to increase the readability of the forest plots, mean values of study groups that were measured by scales where higher values indicate a more positive impact have been reversed (i.e., multiplied by −1).^48^ Cluster-randomized trials were combined with individually randomized controlled trials in the same meta-analyses.^49^ We did not perform author requests for additional or missing data. For all outcome results, we considered the latest follow-up that was reported.

## RESULTS

Our search retrieved a total of 3391 hits. Ninety-three full-text articles were screened against the inclusion criteria. We excluded 80 reports after full-text screening (Figure 1; Appendix 2) and included 9 studies.^16,17,19,21,23–27^ For 3 studies,^16,23,24^ we included additional study reports.^18,20,22,28^ In the following, we no longer refer to these additional reports. Except from two reports that we identified via citation searching methods,^18,26^ we retrieved all reports from database searching. Detailed results for UDCS and RICS will be published elsewhere, citing this report.

**Figure 1.**
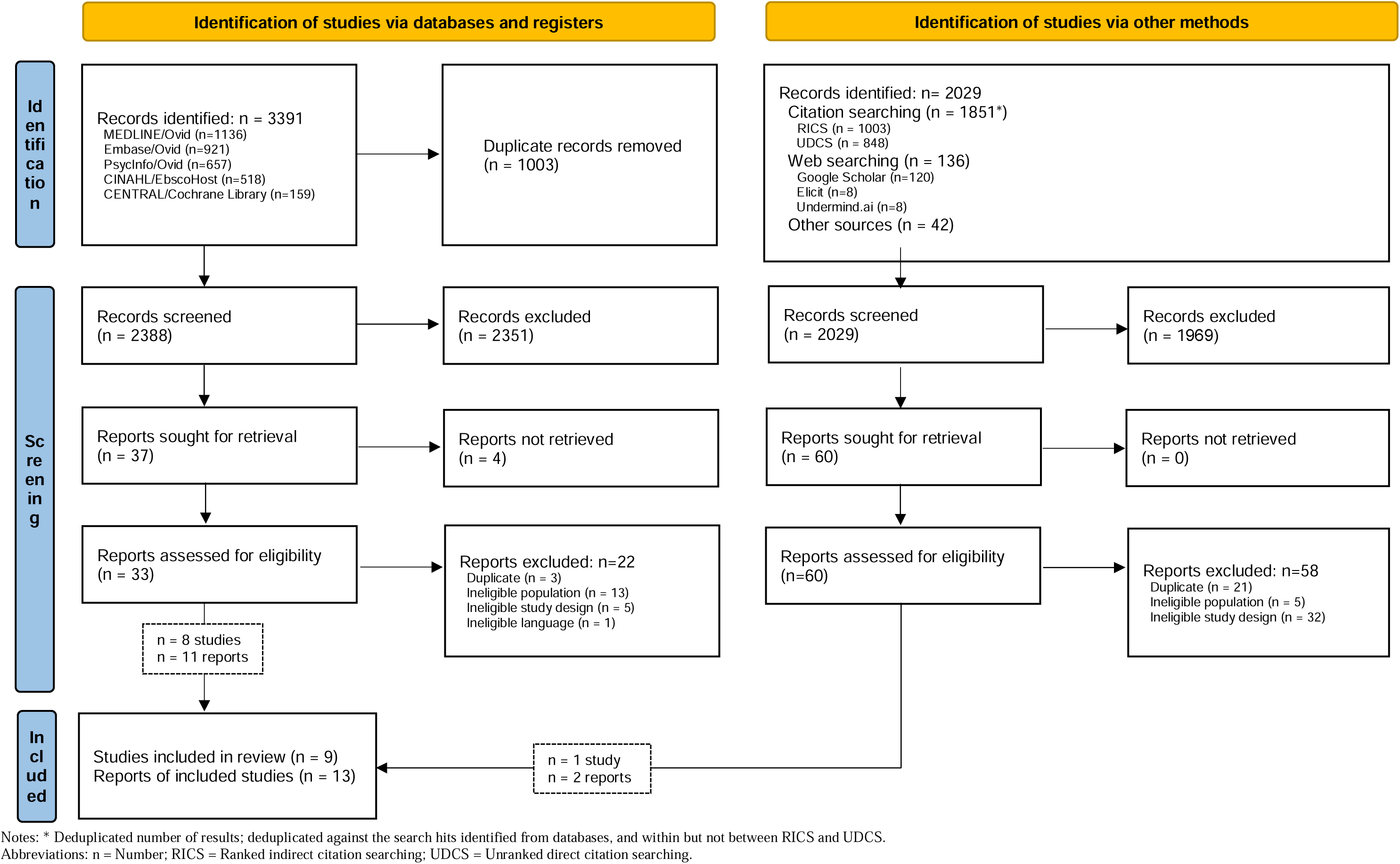
Literature search and selection process

### Characteristics of the included studies

The studies were published between 1990 and 2024 with a median publication year of 2019 (Table 1). Three studies were conducted in the United States (33%),^19,21,26^ 2 in the Netherlands (22%)^16,17^, 2 in Australia (22%),^24,25^ 1 in Italy (11%)^27^, and 1 in the United Kingdom (11%).^23^ Except from 1 study that was a non-randomized study of intervention (NRIS) (11%),^24^ all were randomized controlled trials (RCT) (n=8; 89%). ^16,17,19,21,23,25–27^ In 1 study (11%),^16^ participants were persons living with YOD, 6 studies (67%) included their informal caregivers,^19,21,23,24,26,27^ and 2 studies included dyads (22%).^17,25^ The number of participants ranged between 12 and 274 (median: 58, overall 524). The participants mean age varied between 53 and 64. The type of dementia was characterized as FTD (n=5 studies; 56%)^19,21,23–25^ or YOD (n=4 studies; 44%; Table 1; Table 2; Appendix 3).^16,17,26,27^

**Table 1.** Characteristics of the included studies (n=9)

|  | **n (%)** | **Study reference(s)** |
| --- | --- | --- |
| **Country of conduct** |  |  |
| United States | 3 (33) | 19,21,26 |
| Netherlands | 2 (22) | 16,17 |
| Australia | 2 (22) | 24,25 |
| Italy | 1 (11) | 27 |
| England | 1 (11) | 23 |
| **Study design** |  |  |
| RCT (cluster and individual) | 8 (89) | 16,17,19,21,23,25-27 |
| NSRI | 1 (11) | 24 |
| **Multi-center study** |  |  |
| No | 5 (56) | 16,19,21,24,26 |
| Yes | 3 (33) | 17,23,27 |
| Unclear | 1 (11) | 25 |
| **Publication year** |  |  |
| 1990s | 1 (11) | 26 |
| 2000s | 0 (0) |  |
| 2010s | 5 (56) | 16,19,23-25 |
| 2020s | 3 (33) | 17,21,27 |
| **Availability of study protocol** |  |  |
| No | 8 (89) | 17,19,21,23-27 |
| Yes | 1 (11) | 16 |
| **Availability of registry entry** |  |  |
| No | 5 (56) | 19,21,24,26,27 |
| Yes | 4 (44) | 16,17,23,25 |
| **Type of dementia** |  |  |
| FTD | 5 (56) | 19,21,23-25 |
| YOD | 4 (44) | 16,17,26,27 |
| **Type of participants** |  |  |
| Informal caregivers | 6 (67) | 19,21,23,24,26,27 |
| Persons living with dementia | 1 (11) | 16 |
| Combination of both | 2 (22) | 17,25 |
| **Age of participants** |  |  |
| Mean age <65 years | 9 (100) | 16,17,19,21,23-27 |
| Mean age ≥65 years | 0 (0) | - |
| **Study setting** |  |  |
| Home-based | 6 (67) | 17,21,23-25,27 |
| Nursing home | 2 (22) | 16,26 |
| Combination of both | 1 (11) | 24 |
| **Sample size (median, range, total)** | 24, 12-274, 503 | - |
| **Number of outcomes per study size (median, range, total)** | 5, 3-11, 51 | - |
| **Follow-up length in months (median, range)** | 5, 2.5-18 | - |

Abbreviations: FTD = Frontotemporal dementia; m = Median; n = Number of participants; NSRI = Non-randomized study of intervention; RCT = Randomized controlled trial.; YOD = Young-onset dementia.

**Table 2.**
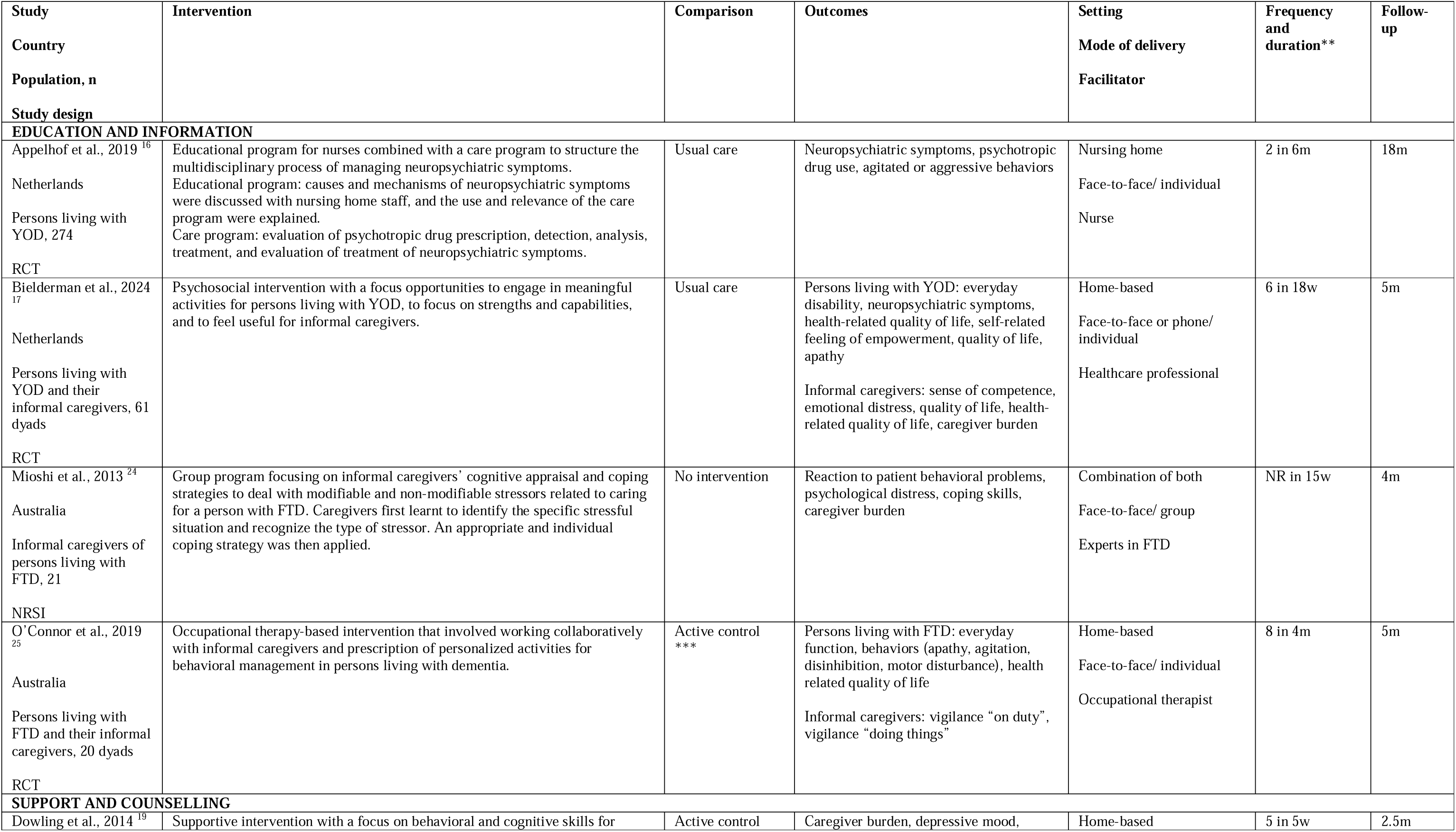
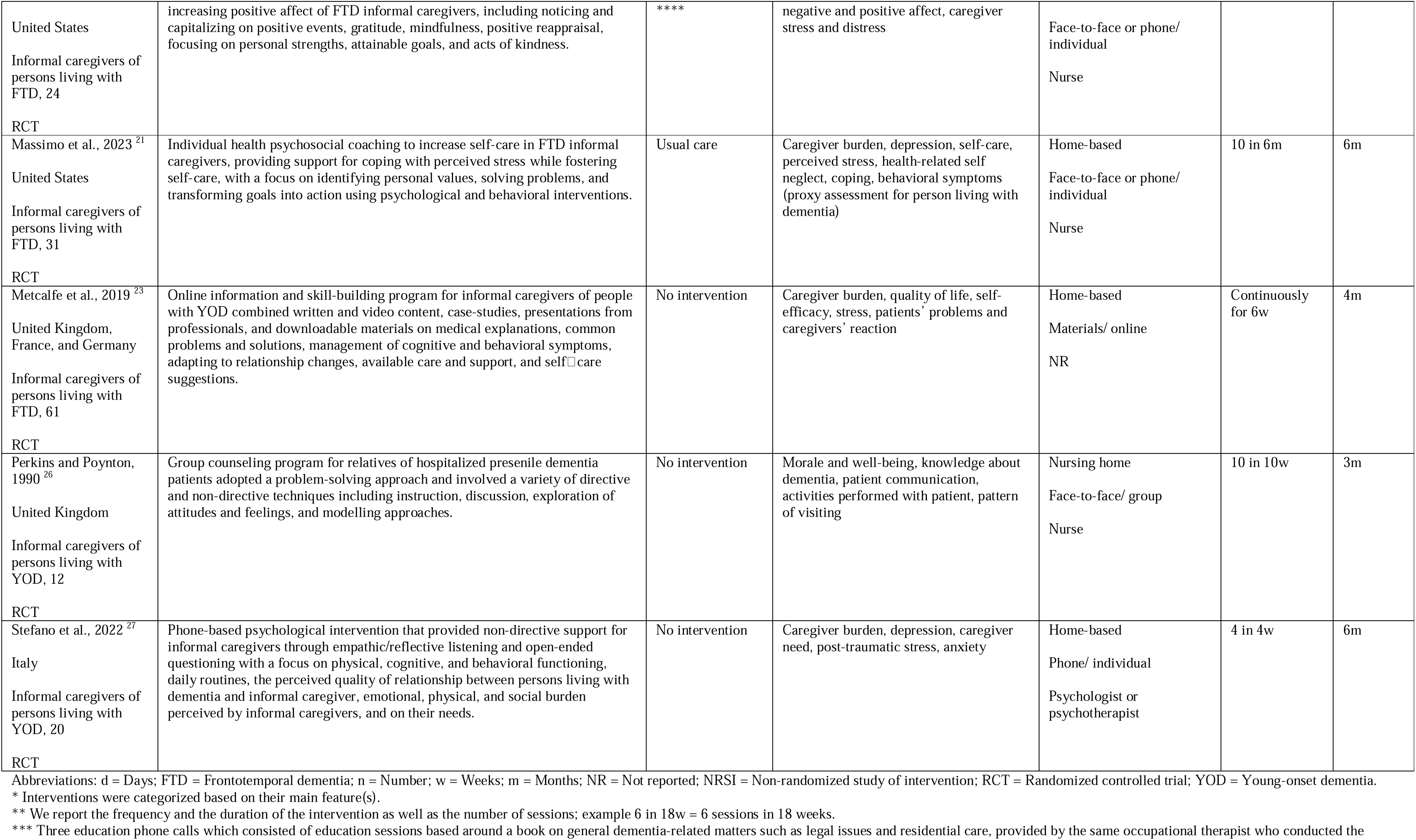
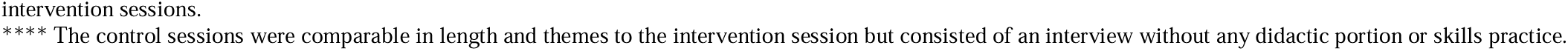
Study and intervention characteristics (n=9)

### Characteristics of the study interventions

The included nine studies assessed interventions that we categorized as education and information (n=4 studies, 44%)^16,17,24,25^ and support and counselling (n=5 studies, 56%).^19,21,23,26,27^ Intervention comparisons were made to usual care (n=3 studies, 33%),^16,17,21^ wait list or no intervention (n=4 studies, 44%)^23,24,26,27^ or active control (n=2 studies, 22%).^19,25^ Six interventions (67%) were delivered individually, either face-to-face (n=2 studies, 22%),^16,25^ phone based (n=1 study, 11%)^27^ or combined (n=3 studies, 33%).^17,19,21^ The other interventions were delivered face-to-face in a group (n= 2 studies, 22%)^24,26^ or with online material (n=1 study, 11%).^23^ The interventions were conducted home-based (n=6 studies, 67%),^17,21,23–25,27^ in nursing homes (n=2 studies, 22%)^16,26^ or in both of them (n=1 study, 11%).^24^ In 4 studies (44%), the intervention was facilitated by nurses,^16,19,21,26^ in 2 studies (22%) by health professionals or experts in dementia care,^17,24^ in 1 study (11%) by occupational therapists,^25^ in 1 study (11%) by psychologists or psychotherapists,^27^ and 1 study (11%) did not report who facilitated the intervention.^23^ The intervention period ranged from 1 month to 6 months. The frequency of the intervention delivery varied between 2 sessions in 6 months to weekly sessions throughout 2.5 months; one study offered the intervention online continuously for 6 weeks (study-specific details in Table 2). The length of follow up ranged from 2.5 to 18 months (Table 2; Appendix 3).

### Methodological quality of included studies

<u>Randomized controlled trials (n=8)</u>: Randomization was appropriately performed in 4 out of 8 studies (50%). Groups were comparable at baseline in 7 out of 8 studies (88%). Outcome data were completed in 6 out of 8 studies (75%). In 5 out of 8 studies (67%), it was unclear if the outcome assessors were blinded to the intervention provided. In 6 out of 8 studies (75%), it was unclear if the participants did adhere to the assigned intervention (Table 3; Appendix 3).

**Table 3.** Methodological quality of the included studies (n=9)

| **RCT** | **Is the randomization appropriately performed?** | **Are the groups comparable at baseline?** | **Are there completed outcome data?** | **Are the outcome assessors blinded to the intervention provided?** | **Did the participants adhere to the assigned intervention?** |
| --- | --- | --- | --- | --- | --- |
| Appelhof et al., 2019 <sup>16</sup> | Can't tell | Can't tell | No | Can't tell | Yes |
| Bielderman et al., 2024 <sup>17</sup> | Yes | Yes | Yes | Can't tell | Yes |
| Dowling et al., 2014 <sup>19</sup> | Yes | Yes | Yes | Can't tell | Can't tell |
| Massimo et al., 2023 <sup>21</sup> | Yes | Yes | Yes | Yes | Can't tell |
| Metcalfe et al., 2019 <sup>23</sup> | Yes | Yes | Yes | Yes | Can't tell |
| O'Connor et al., 2019 <sup>25</sup> | Yes | Yes | Yes | Yes | Can't tell |
| Perkins and Poynton, 1990 <sup>26</sup> | Can't tell | Yes | Can't tell | Can't tell | Can't tell |
| Stefano et al., 2022 <sup>27</sup> | Can't tell | Yes | Yes | Can't tell | Can't tell |
| **NRSI** | **Are the participants representative of the target population?** | **Are measurement instruments appropriate regarding both the outcome and intervention (or exposure)?** | **Are there complete outcome data?** | **Are the confounders accounted for in the design and analysis?** | **During the study period, is the intervention administered (or exposure occurred) as intended?** |
| Mioshi et al., 2013 <sup>24</sup> | Yes | Yes | Can't tell | Yes | Yes |

Abbreviations: NRSI = Non-randomized study of intervention; RCT = Randomized controlled trial.

<u>Non-randomized studies of intervention (n=1)</u>: In this single study, participants were assessed as being representative of the target population. Measurement instruments were appropriate regarding both outcome and intervention. It is unclear if there were complete outcome data due to insufficient reporting. Confounders were accounted for the design and analysis. The intervention was administered as intended during the study period (Table 3; Appendix 3).

### Qualitative synthesis of results

Eight studies that assessed the effects on 10 outcomes were considered for qualitative/narrative synthesis as shown in the harvest plot (n = 8 studies; follow-up range between 1 and 18 months, median = 14 months; Figure 2) and summarized below.

**Figure 2.**
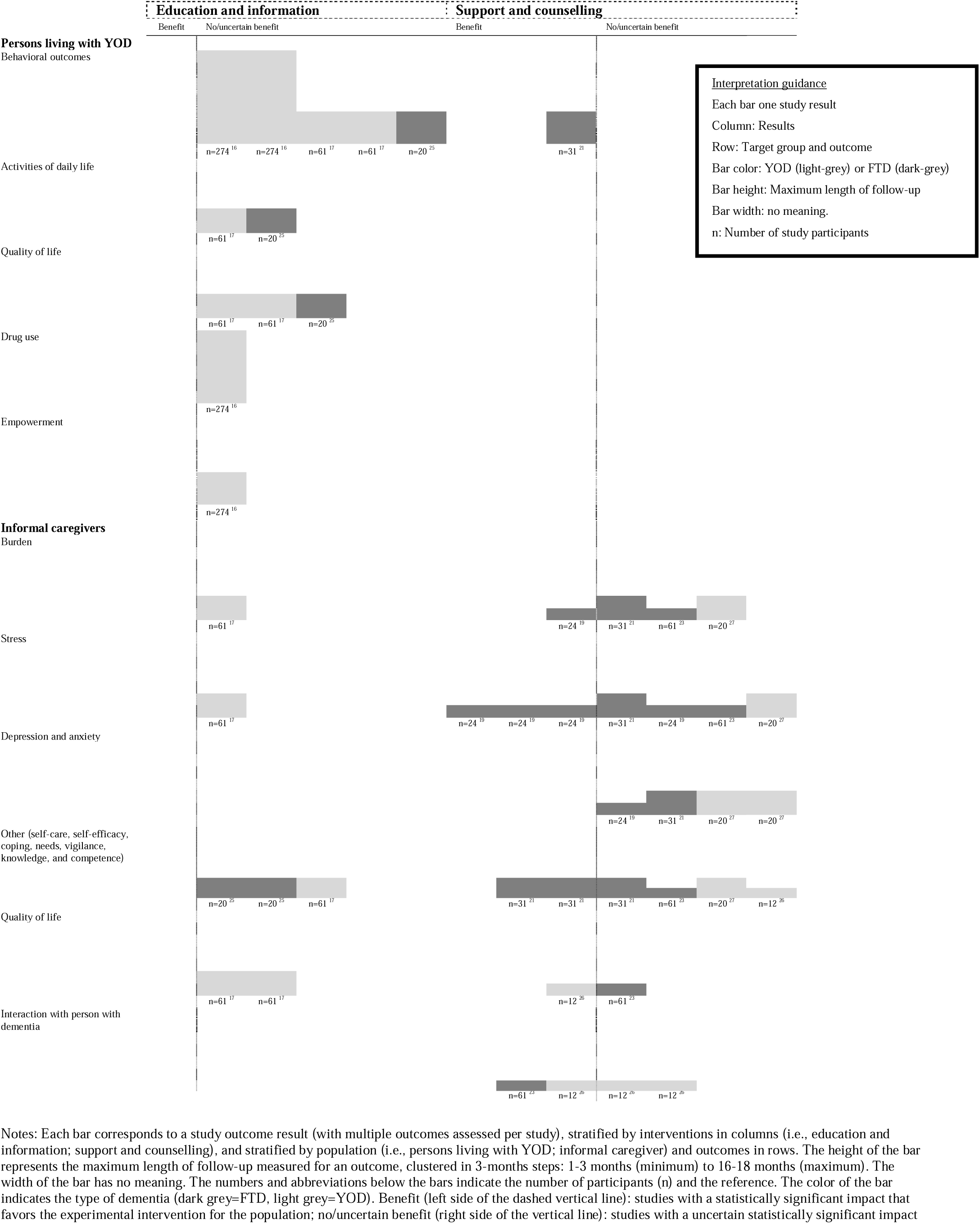
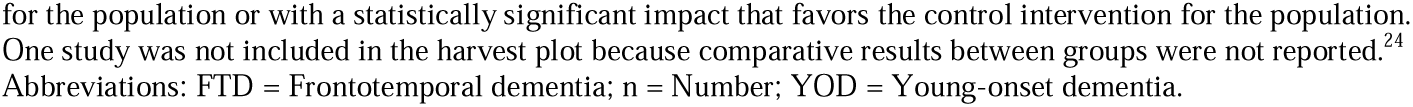
Harvest plot with effect directions of study outcomes and results

<u>Education and information for persons living with YOD</u>: No statistically significant beneficial impact on behavioral outcomes (i.e., neuropsychiatric symptoms or single behavioral symptoms including apathy, agitation, aggression, disinhibition, motor disturbance; Appendix 3),^16,17,25^ activities of daily living,^17,25^ quality of life,^17,25^ drug use^16^ or empowerment.^16^

Support and counselling for persons living with YOD: Support and counselling proved to have a statistically significant positive impact on behavioral outcomes.^21^

Education and information for informal caregivers: No statistically significant beneficial impact on burden,^17^ stress,^17^ quality of life,^17^ and other outcomes (self-care, self-efficacy, coping, needs, vigilance, knowledge and competence).^17,25^

For one study in which comparative results between groups were not reported, within group results indicated a statistically significant decrease in the level of informal caregiver burden and reaction of behaviors in the intervention group immediately after the intervention compared to baseline values (with no statistically significant changes on depression, anxiety, and stress; and no statistically significant within group changes on any outcome in the control group).^24^

<u>Support and counselling for informal caregivers</u>: No statistically significant beneficial impact on depression and anxiety.^19,21,27^ For burden and stress, one study reported significant benefits,^19^ while others found no statistically significant effect.^19,21,23,27^ For quality of life, one study each showed a beneficial^26^ and no beneficial^23^ statistically significant impact. Two studies showed significant positive impact on interaction with persons living with YOD,^23,26^ though one also showed no statistically significant benefit in other domains in this category.^26^ Other outcomes (self-care, self-efficacy, coping, needs, vigilance, knowledge and competence) showed both statistically significant ^21^ and insignificant beneficial impact.^21,23,26,27^

### Quantitative synthesis of results

Eight studies that assessed the effects on 6 outcomes were considered for quantitative/statistical analysis (Figure 3; Figure 4) and summarized below.

**Figure 3.**
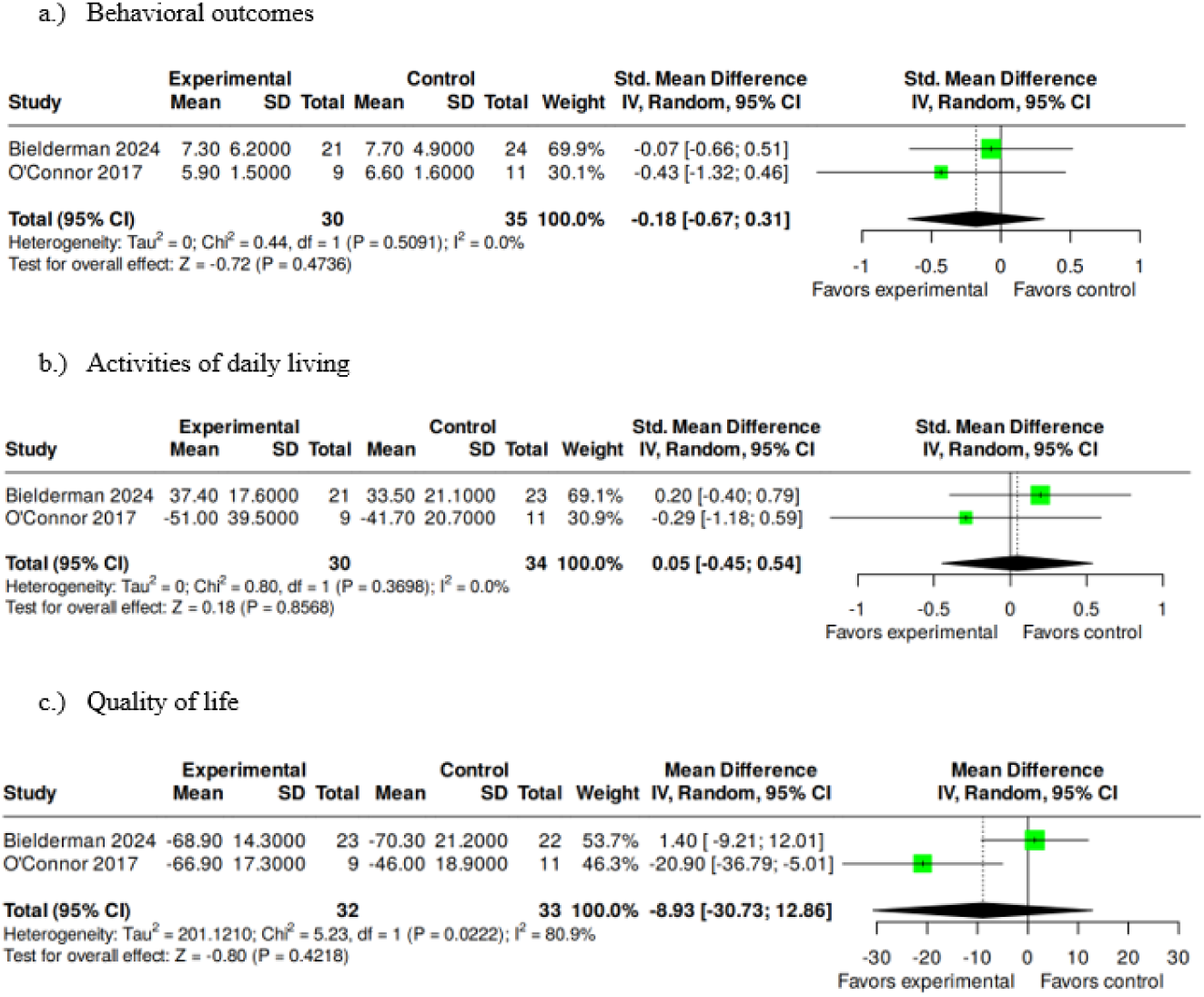
Forest plot on education and information interventions for persons living with YOD

**Figure 4.**
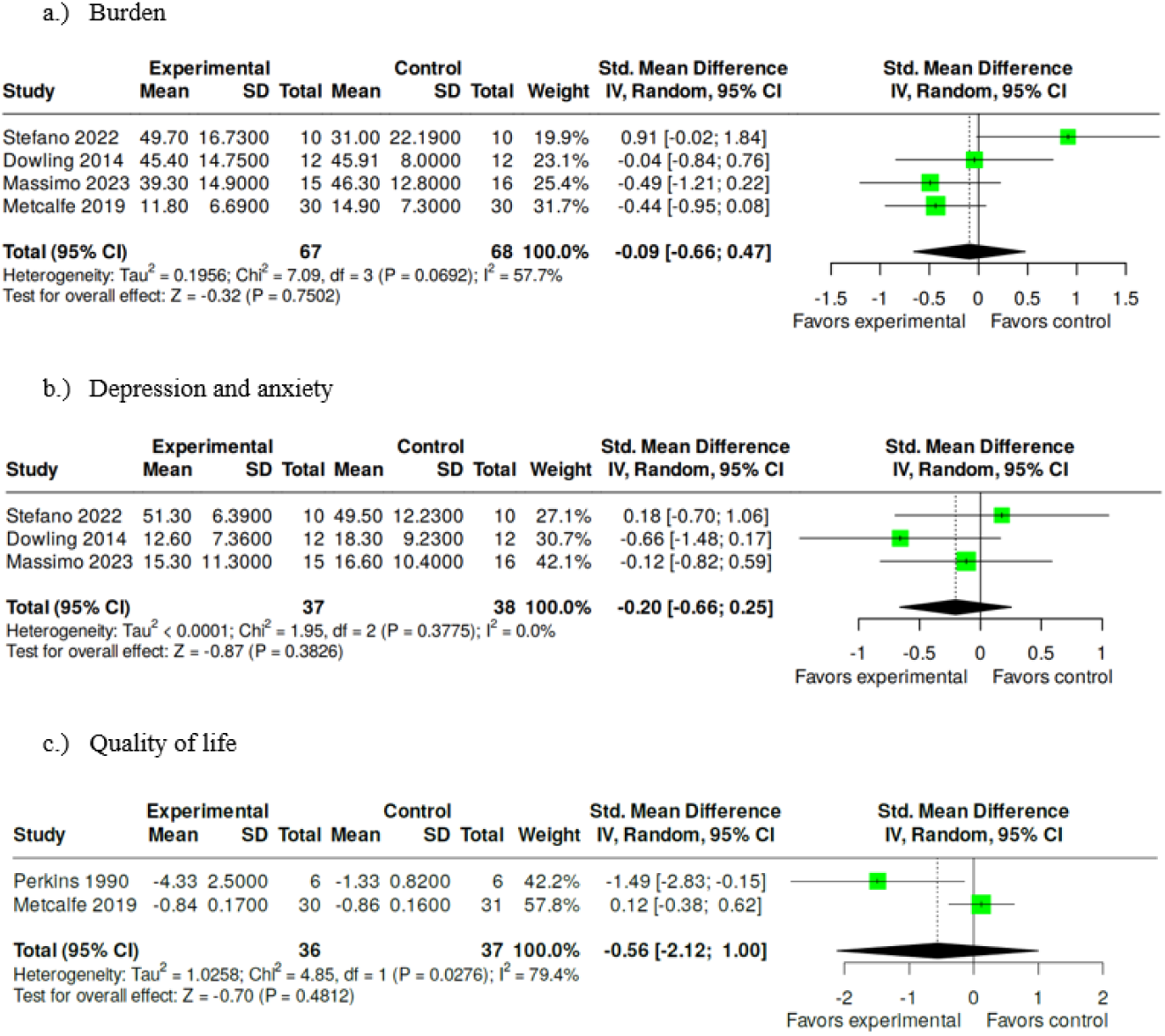
Forest plot on support and counselling interventions for informal caregivers

Education and information for persons living with YOD: There were no statistical group differences on behavioral outcomes (SMD −0.18, 95% CI −0.67; 0.31),^17,25^ activities of daily living (SMD 0.05, 95% CI −0.45; 0.54),^17,25^ and quality of life (MD −8.93, 95% CI −30.73; 12.86).^17,25^

Support and counselling for informal caregivers: There were no statistical group differences on burden (SMD −0.09, 95% CI −0.66; 0.47),^19,21,23,27^ depression and anxiety (SMD −0.20, 95% CI −0.66; 0.25),^19,21,27^ and quality of life (SMD −0.56, 95% CI −2.12; 1.00).^23,26^

## DISCUSSION

We included eight studies assessing the impact of non-pharmaceutical interventions for persons living with YOD and their informal caregivers. Educational and informational interventions for persons living with YOD had no statistically significant impact on behavioral outcomes, daily activities, quality of life, drug use, or empowerment. Support and counselling showed a positive effect on behavioral outcomes in one study. For informal caregivers, education and information had no statistically significant effects, while support and counselling showed mixed results, with some benefits in burden, quality of life, and interaction. The statistical synthesis widely confirmed the narrative findings. No statistically significant differences were observed between intervention and control groups for persons living with YOD in behavior, activities of daily living, or quality of life. Similarly, among informal caregivers, there were no statistically significant effects of support and counselling on burden, depression and anxiety, or quality of life.

### Impact of interventions for persons living with YOD

There was a statistically significant beneficial effect on behavioral symptoms in persons living with FTD in one small study with 31 participants in which informal caregivers of persons living with FTD received individual health psychosocial coaching.^21^ A narrative review identified interventions that also aimed to assist informal caregivers in learning strategies to positively impact behavioral outcomes, e.g. by teaching how to respond and modify the behavioral response of the person with FTD over time, meeting unmet needs of the individuals or focusing on the role that their own and environmental status can play as triggers for behavioral symptoms. Thus, these interventions may reduce behavioral symptoms, such as agitation and aggression in dementia populations.^37^

Education and information interventions showed no statistically beneficial impact on behavioral outcomes, activities of daily living, quality of life, drug use and empowerment.^16,17,25^ Even if information is known as an unmet need of people with YOD^7^ and qualitative studies that evaluated psycho-educational interventions showed that participants generally expressed positive satisfaction with the interventions,^7^ the results of our review suggest that education and information interventions might not have a positive impact. Non-pharmaceutical interventions such as cognitive rehabilitations, physical and social activities, psychosocial interventions, and environmental support showed partial improvements for persons living with YOD and positive satisfaction with the interventions. These findings, however, were derived from both controlled and non-controlled designs, unlike our review, which included only controlled trials.^7^ Even if interventions included in our review did not show statistically beneficial impact on most outcomes, considering qualitative results, non-pharmaceutical interventions were experienced positively as they provided persons living with YOD a sense of purpose, enhanced their happiness, improved their ability to engage in independent activities, establish goals, and uplifted their mood.^10^

### Impact of interventions for informal caregivers of persons living with YOD

There were no statistically significant beneficial impacts when assessing education and information interventions.^17,25^ Our qualitative synthesis showed mixed benefits of support and counselling interventions on burden, stress, quality of life, interaction and others (e.g., knowledge, self-care) and no benefit on depression and anxiety.^19,21,23,26,27^ Meta-analyses showed no statistically significant benefits on burden, quality of life, and depression and anxiety.^19,21,23,26,27^

Taken together, these findings suggest that while support and counselling approaches may offer some benefits, their effects remain inconsistent across studies and outcomes. This variability has prompted growing interest in alternative modes of delivery, particularly technology-based interventions.^36,50^ In our review, we included one technology-based intervention, which provided informal caregivers of people with FTD with an online information and skill-building program. The program combined written and video content, case studies, and presentations from professionals. It also offered downloadable materials on medical explanations, common problems and solutions, the management of cognitive and behavioral symptoms, adapting to relationship changes, available care and support, and suggestions for self-care.^23^ Based on 12 qualitative and quantitative (of which 3 RCTs) studies conducted in Europe and Canada, Moyle et al. (2025) identified technologies that support informal caregivers of persons living with YOD. These technologies included online support interventions with information and skill-building programs, virtual psychotherapy, telephone-based psychological intervention, assistive technologies such as sensor and location devices, automated timers, mobile phone, automatic calendar, talking wristwatch, a medicine dispenser, a TV remote control, and telehealth. The interventions showed a positive impact on self-efficacy, satisfaction, knowledge, well-being, perceived burden and stress, depression and anxiety, except for one study which did not find any significant differences in measured outcomes. In addition, informal caregivers rated the program overall positively, demonstrating high satisfaction and acceptability.^35^ Another scoping review identified web-based and face-to-face intervention studies that supported informal caregivers of persons living with YOD in managing the changes brought about by the disease showed significant but also no or unclear significant benefits of these interventions. However, qualitative findings indicated positive changes, including improved well-being, confidence in coping with challenges, and transition process but challenges were also identified in the scoping review, including technological difficulties.^10^ The positive effects of technological interventions could be attributed to their ease of integration into daily life and the relative anonymity they offer, which facilitates the expression of true feelings and experiences. They also save logistical workforce resources compared to traditional face-to-face interventions.^10^

### Characteristics of interventions

The identified interventions differed in terms of type, content, format, setting, and duration. They were delivered individually or in a group, face-to-face, phone-based or online in nursing homes or home-based. The facilitators were health professionals, most of them nurses. The duration varied widely. Some interventions focused solely on providing information, whereas others combined information with counselling or skills training. Importantly, interventions that relied on information and education alone appeared insufficient to produce significant benefits, suggesting that information may need to be embedded within broader frameworks. In their scoping review, Cui et al. (2024) focused on non-pharmaceutical interventions for persons living with YOD and/or their informal caregivers and also identified a variety of interventions in terms of duration, frequency, content, and delivery. The interventions were delivered by different health professionals and were offered individually or group-based in different settings, such as remote, nursing-homes, home-based or in zoos, gardens, or football clubs. The authors argued that information alone often fall short in achieving meaningful impact, and should be combined with other components like skills training or psychological support.^10^ Also Kim et al. (2024) emphasized the need for combined approaches, rather than single-focus interventions, as being more aligned with the complex needs of persons living with YOD and their informal caregivers. ^7^

Our review is based on nine studies with a total sample size of 503 participants with a median of 24. Other reviews on the topic also reported small numbers of studies participants with mostly short lengths of follow-up.^7,10,11,35,36^ Additionally, our review includes only two studies with dyads of persons living with YOD and their informal caregivers, indicating a need for intervention studies that address the entire family of a person living with YOD. It is necessary that interventions understand and tailor on the care context of persons living with YOD and their informal caregivers to consider their needs and enhance quality of life.^7,36^ To do so, persons living with YOD and their informal caregivers can be included in the design of support groups or the project steering group or committee.^36^

### Limitations

The strength of this review is a comprehensive search strategy considering databases, direct and indirect citation searching, and web- and ai-based searches. Study selection by two independent reviewers with clinical and methodological expertise and verification of data extraction and synthesis served to ensure high data quality. Although we did not plan to contact study authors for missing outcome data, we were able to run all meaningful meta-analyses. Our review also has some limitations. First, our search missed the term ‘familial Alzheimer disease’ and ‘presenile dementia’. However, we explored the related search hits on PubMed and do not believe that we missed eligible study reports. Second, we did not perform a robust risk of bias assessment of the study results. This, in combination with a certainty assessment of the evidence, should be considered in future assessments when aiming to derive clinical implications of the evidence; which was not the aim of our review. Finally, we did not perform a detailed assessment of the study intervention. Applying assessments such as the ‘Template for Intervention Description and Replication’ (TIDieR) checklist, ^51^ the updated ‘Criteria for Reporting the Development and Evaluation of Complex Interventions’ (CReDECI-2) guideline,^52^ or the more recent ‘Guidance for reporting intervention development studies in health research’ (GUIDED)^53^ may help to understand the interventions’ mechanisms and plan future interventions for persons living with YOD and their informal caregivers.

## CONCLUSIONS

Evidence on the effectiveness of non-pharmaceutical interventions for persons living with YOD and their informal caregivers is limited and inconsistent, highlighting that most of the studies are very small with short follow-ups, heterogenous interventions and outcomes, and that there is a lack of studies on interventions targeting both persons living with YOD and their informal caregivers. The non-pharmacological interventions studied were primarily educational, informational, and supportive or consulting in nature. Educational and informational interventions showed no significant impact, while support and counselling interventions yielded mixed results. Statistical analyses confirmed the absence of robust effects. To increase the quality and certainty of evidence on non-pharmaceutical interventions for persons living with YOD and their informal caregivers, further, larger, and multiple randomized controlled trials assessing the impact of specific non-pharmaceutical interventions with comparable outcomes, standardized measurements, and longer follow-ups are needed.

## DECLARATIONS

### Funding

This review was part of an overarching project focusing on persons living with young-onset dementia aiming at developing a concept to accompany and support persons living with young-onset and frontotemporal dementia and their families, funded by Gesundheitsförderung Schweiz (PortoFaro; https://gesundheitsfoerderung.ch/praevention-in-der-gesundheitsversorgung/projektfoerderung/gefoerderte-projekte/portofaro). The funder had no influence on the design, conduct, and reporting of this review.

### Patient and public involvement

This review is part of the PortoFaro project which has been planned in close collaboration with patient and public representatives (https://gesundheitsfoerderung.ch/praevention-in-der-gesundheitsversorgung/projektfoerderung/gefoerderte-projekte/portofaro).

### Competing interests

None to declare.

### Ethics approval and consent to participate

Not applicable.

### Consent for publication

Not applicable.

### Availability of data and material

All data generated and analyzed in this study is part of the publication and appendix.

Detailed results for the unranked direct citation search and ranked indirect citation search will be published elsewhere, citing this report.

## Acknowledgement

We thank Cristina De Biasio and Linda Premerlani (both mosa!k St.Gallen and PortoFaro project team members) for contributing to the design of this review (i.e., review questions and eligibility criteria).

## CRediT (Contributor Roles Taxonomy) author statement

Magdalena Vogt: Methodology, Validation, Formal analysis, Investigation, Writing - Original Draft, Writing - Review & Editing, Visualization

Nicole Helfenberger: Validation, Investigation, Writing - Review & Editing

Christian Appenzeller-Herzog: Methodology, Writing - Review & Editing, Supervision

Laura Adlbrecht: Conceptualization, Methodology, Writing - Review & Editing, Project administration, Funding acquisition

Julian Hirt: Conceptualization, Methodology, Software, Validation, Formal analysis, Investigation, Writing −

Original Draft, Writing - Review & Editing, Visualization, Supervision, Project administration, Funding acquisition

## Appendix 1. Database-specific search strategies

**MEDLINE/Ovid**

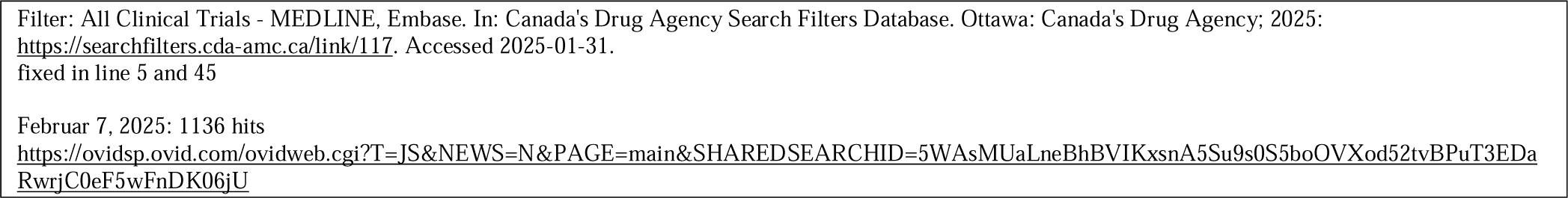

((“young onset” OR “younger onset” OR “early onset” OR “young* people” OR “young* person*” OR frontotemporal).ti,ab,hw,kf. adj3 (“DEMENT*” OR “ALZHEIMER*”).ti,ab,hw,kf.) OR (exp Frontotemporal Dementia/ OR FTD.ti,ab,hw,kf. OR “pick* disease”.ti,ab,hw,kf. OR “Wilhelmsen* Disease”.ti,ab,hw,kf.)

AND

(((Randomized Controlled Trial or Controlled Clinical Trial or Pragmatic Clinical Trial or Clinical Study or Adaptive Clinical Trial or Equivalence Trial).pt. or (Clinical Trial or Clinical Trial, Phase I or Clinical Trial, Phase II or Clinical Trial, Phase III or Clinical Trial, Phase IV or Clinical Trial Protocol).pt. or Multicenter Study.pt. or
Clinical Studies as Topic/ or
exp Clinical Trial/ or exp Clinical Trials as Topic/ or Clinical Trial Protocol/ or Clinical Trial Protocols as Topic/ or “Clinical Trial (topic)”/ or Multicenter Study/ or Multicenter Studies as Topic/ or “Multicenter Study (topic)”/ or
Randomization/ or
Random Allocation/ or
Double-Blind Method/ or
Double Blind Procedure/ or
Double-Blind Studies/ or
Single-Blind Method/ or
Single Blind Procedure/ or
Single-Blind Studies/ or
Placebos/ or
Placebo/ or
Control Groups/ or
Control Group/ or
Cross-Over Studies/ or Crossover Procedure/ or
(random* or sham or placebo*).ti,ab,hw,kf. or
((singl* or doubl*) adj (blind* or dumm* or mask*)).ti,ab,hw,kf. or
((tripl* or trebl*) adj (blind* or dumm* or mask*)).ti,ab,hw,kf. or
(control* adj3 (study or studies or trial* or group*)).ti,ab,hw,kf. or
(clinical adj3 (study or studies or trial*)).ti,ab,hw,kf. or
(Nonrandom* or non random* or non-random* or quasi-random* or quasirandom*).ti,ab,hw,kf. or
(phase adj6 (study or studies or trial*)).ti,ab,hw,kf. or
((crossover or cross-over) adj3 (study or studies or trial*)).ti,ab,hw,kf. or
((multicent* or multi-cent*) adj3 (study or studies or trial*)).ti,ab,hw,kf. or
allocated.ti,ab,hw. Or
((open label or open-label) adj5 (study or studies or trial*)).ti,ab,hw,kf. or
((equivalence or superiority or non-inferiority or noninferiority) adj3 (study or studies or trial*)).ti,ab,hw,kf. or
(pragmatic study or pragmatic studies).ti,ab,hw,kf. or
((pragmatic or practical) adj3 trial*).ti,ab,hw,kf. or
((quasiexperimental or quasi-experimental) adj3 (study or studies or trial*)).ti,ab,hw,kf. or trial.ti,kf.)
not
((exp animals/ or
exp animal experimentation/ or
exp models animal/ or
exp animal experiment/ or
nonhuman/ or
exp vertebrate/)
not
(exp humans/ or
exp human experimentation/)))

NOT

(genetic*.ti,ab,hw,kf. OR tau.ti,ab,hw,kf. OR amyloid*.ti,ab,hw,kf. OR SURGICAL*.ti,ab,hw,kf. OR SURGERY*.ti,ab,hw,kf. OR PROTEOMIC*.ti,ab,hw,kf. OR GENOMIC*.ti,ab,hw,kf. OR PROTEIN*.ti,ab,hw,kf. OR biomarker*.ti,ab,hw,kf. OR Presenilin*.ti,ab,hw,kf. OR Progranulins.ti,ab,hw,kf. OR peptid*.ti,ab,hw,kf. OR “Niemann-Pick”.ti,ab,hw,kf.)

**Embase/Ovid**

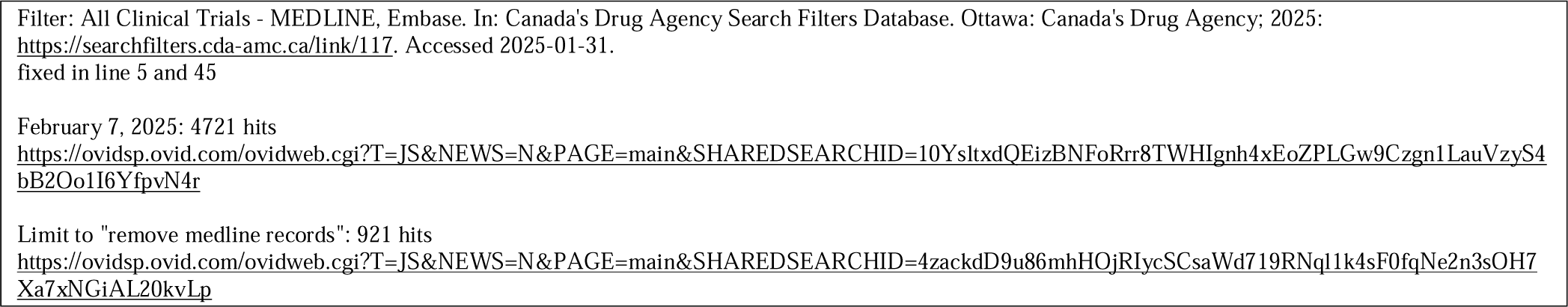

((“young onset” OR “younger onset” OR “early onset” OR “young* people” OR “young* person*” OR frontotemporal).ti,ab,hw,kf. adj3 (“DEMENT*” OR “ALZHEIMER*”).ti,ab,hw,kf.) OR (exp Frontotemporal Dementia/ OR FTD.ti,ab,hw,kf. OR “pick* disease”.ti,ab,hw,kf. OR “Wilhelmsen* Disease”.ti,ab,hw,kf.)

AND

(((Randomized Controlled Trial or Controlled Clinical Trial or Pragmatic Clinical Trial or Clinical Study or Adaptive Clinical Trial or Equivalence Trial).pt. or (Clinical Trial or Clinical Trial, Phase I or Clinical Trial, Phase II or Clinical Trial, Phase III or Clinical Trial, Phase IV or Clinical Trial Protocol).pt. or Multicenter Study.pt. or
Clinical Studies as Topic/ or
exp Clinical Trial/ or exp Clinical Trials as Topic/ or Clinical Trial Protocol/ or Clinical Trial Protocols as Topic/ or “Clinical Trial (topic)”/ or Multicenter Study/ or Multicenter Studies as Topic/ or “Multicenter Study (topic)”/ or
Randomization/ or Random Allocation/ or Double-Blind Method/ or Double Blind Procedure/ or Double-Blind Studies/ or
Single-Blind Method/ or
Single Blind Procedure/ or
Single-Blind Studies/ or
Placebos/ or
Placebo/ or
Control Groups/ or
Control Group/ or
Cross-Over Studies/ or Crossover Procedure/ or
(random* or sham or placebo*).ti,ab,hw,kf. or
((singl* or doubl*) adj (blind* or dumm* or mask*)).ti,ab,hw,kf. or
((tripl* or trebl*) adj (blind* or dumm* or mask*)).ti,ab,hw,kf. or
(control* adj3 (study or studies or trial* or group*)).ti,ab,hw,kf. or
(clinical adj3 (study or studies or trial*)).ti,ab,hw,kf. or
(Nonrandom* or non random* or non-random* or quasi-random* or quasirandom*).ti,ab,hw,kf. or
(phase adj6 (study or studies or trial*)).ti,ab,hw,kf. or
((crossover or cross-over) adj3 (study or studies or trial*)).ti,ab,hw,kf. or
((multicent* or multi-cent*) adj3 (study or studies or trial*)).ti,ab,hw,kf. or
allocated.ti,ab,hw. Or
((open label or open-label) adj5 (study or studies or trial*)).ti,ab,hw,kf. or
((equivalence or superiority or non-inferiority or noninferiority) adj3 (study or studies or trial*)).ti,ab,hw,kf. or
(pragmatic study or pragmatic studies).ti,ab,hw,kf. or
((pragmatic or practical) adj3 trial*).ti,ab,hw,kf. or
((quasiexperimental or quasi-experimental) adj3 (study or studies or trial*)).ti,ab,hw,kf. or
trial.ti,kf.)
not
((exp animals/ or
exp animal experimentation/ or
exp models animal/ or
exp animal experiment/ or
nonhuman/ or
exp vertebrate/) not
(exp humans/ or
exp human experimentation/)))

NOT

(genetic*.ti,ab,hw,kf. OR tau.ti,ab,hw,kf. OR amyloid*.ti,ab,hw,kf. OR SURGICAL*.ti,ab,hw,kf. OR SURGERY*.ti,ab,hw,kf. OR PROTEOMIC*.ti,ab,hw,kf. OR GENOMIC*.ti,ab,hw,kf. OR PROTEIN*.ti,ab,hw,kf. OR biomarker*.ti,ab,hw,kf. OR Presenilin*.ti,ab,hw,kf. OR Progranulins.ti,ab,hw,kf. OR peptid*.ti,ab,hw,kf. OR “Niemann-Pick”.ti,ab,hw,kf.)

NOT

Conference abstract.pt

**PsycInfo/Ovid**

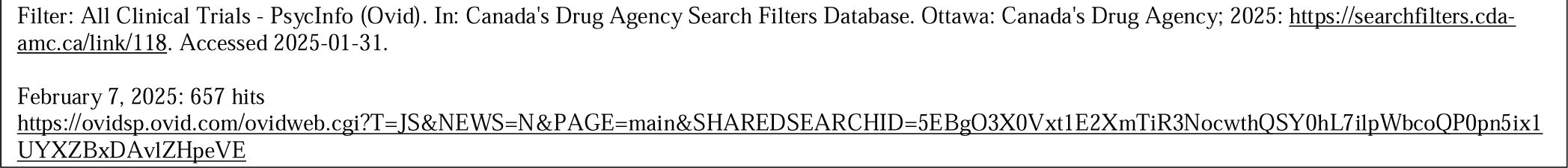

((“young onset” OR “younger onset” OR “early onset” OR “young* people” OR “young* person*” OR frontotemporal).ti,ab,id,hw,mf. adj3 (“DEMENT*” OR “ALZHEIMER*”).ti,ab,id,hw,mf.) OR (exp Frontotemporal Dementia/ OR FTD.ti,ab,id,hw,mf. OR “pick* disease”.ti,ab,id,hw,mf. OR “Wilhelmsen* Disease”.ti,ab,id,hw,mf.)

AND

(Experiment Controls/ or Placebo/ or Clinical Trials/ or Randomized Controlled Trials/ or Randomized Clinical Trials/ or Clinical trial.md. or
(random* or sham or placebo*).ti,ab,id,hw,mf. or
((singl* or doubl*) adj (blind* or dumm* or mask*)).ti,ab,id,hw,mf. or
((tripl* or trebl*) adj (blind* or dumm* or mask*)).ti,ab,id,hw,mf. or
(control* adj3 (study or studies or trial* or group*)).ti,ab,id,hw,mf. or
(clinical adj3 (study or studies or trial*)).ti,ab,id,hw,mf. or
(Nonrandom* or non random* or non-random* or quasi-random* or quasirandom*).ti,ab,id,hw,mf. or
(phase adj6 (study or studies or trial*)).ti,ab,id,hw,mf. or
((crossover or cross-over) adj3 (study or studies or trial*)).ti,ab,id,hw,mf. or
((multicent* or multi-cent*) adj3 (study or studies or trial*)).ti,ab,id,hw,mf. or
allocated.ti,ab,hw,mf. or
((open label or open-label) adj5 (study or studies or trial*)).ti,ab,id,hw,mf. or
((equivalence or superiority or non-inferiority or noninferiority) adj3 (study or studies or trial*)).ti,ab,id,hw,mf. or
(pragmatic study or pragmatic studies).ti,ab,id,hw,mf. or
((pragmatic or practical) adj3 trial*).ti,ab,id,hw,mf. or
((quasiexperimental or quasi-experimental) adj3 (study or studies or trial*)).ti,ab,id,hw,mf. or
trial.ti,id.)

NOT

(genetic*.ti,ab,id,hw,mf. OR tau.ti,ab,id,hw,mf. OR amyloid*.ti,ab,id,hw,mf. OR SURGICAL*.ti,ab,id,hw,mf. OR SURGERY*.ti,ab,id,hw,mf. OR PROTEOMIC*.ti,ab,id,hw,mf. OR GENOMIC*.ti,ab,id,hw,mf. OR PROTEIN*.ti,ab,id,hw,mf. OR biomarker*.ti,ab,id,hw,mf. OR Presenilin*.ti,ab,id,hw,mf. OR Progranulins.ti,ab,id,hw,mf. OR peptid*.ti,ab,id,hw,mf. OR “Niemann-Pick”.ti,ab,id,hw,mf.)

**CINAHL/EbscoHost**

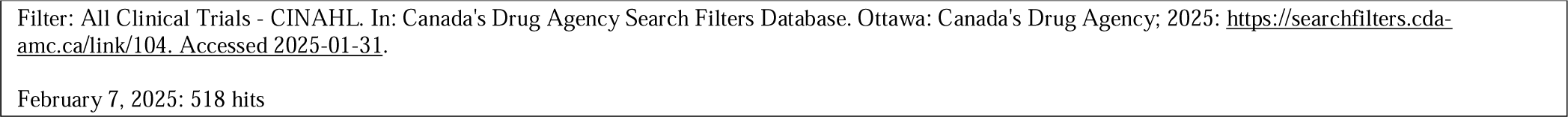

((((TI “young onset” OR AB “young onset”) OR (TI “younger onset” OR AB “younger onset”) OR (TI “early onset” OR AB “early onset”) OR (TI “young* people” OR AB “young* people”) OR (TI “young* person*” OR AB “young* person*”) OR (TI frontotemporal OR AB frontotemporal)) N3 ((TI DEMENT* OR AB DEMENT*) OR (TI ALZHEIMER* OR AB ALZHEIMER*))) OR ((MH “Frontotemporal Dementia+”) OR (TI FTD OR AB FTD) OR (TI “pick* disease” OR AB “pick* disease”) OR (TI “Wilhelmsen* Disease” OR AB “Wilhelmsen* Disease”)))

AND

(((MH “Experimental Studies+”) OR (MH “Multicenter Studies”) OR (MH “Random Sample+”) OR (MH “Placebos”) OR (MH “Control (Research)+”) OR (MH “Crossover Design”) OR ((TI random* OR AB random*) OR (TI sham OR AB sham) OR (TI placebo* OR AB placebo*)) OR (((TI singl* OR AB singl*) OR (TI doubl* OR AB doubl*)) W1 ((TI blind* OR AB blind*) OR (TI dumm* OR AB dumm*) OR (TI mask* OR AB mask*))) OR (((TI tripl* OR AB tripl*) OR (TI trebl* OR AB trebl*)) W1 ((TI blind* OR AB blind*) OR (TI dumm* OR AB dumm*) OR (TI mask* OR AB mask*))) OR ((TI control* OR AB control*) N3 ((TI study OR AB study) OR (TI studies OR AB studies) OR (TI trial* OR AB trial*) OR (TI group* OR AB group*))) OR ((TI clinical OR AB clinical) N3 ((TI study OR AB study) OR (TI studies OR AB studies) OR (TI trial* OR AB trial*))) OR ((TI Nonrandom* OR AB Nonrandom*) OR (TI “non random*” OR AB “non random*”) OR (TI “non-random*” OR AB “non-random*”) OR (TI “quasi-random*” OR AB “quasi-random*”) OR (TI quasirandom* OR AB quasirandom*)) OR ((TI phase OR AB phase) N6 ((TI study OR AB study) OR (TI studies OR AB studies) OR (TI trial* OR AB trial*))) OR (((TI crossover OR AB crossover) OR (TI “cross-over” OR AB “cross-over”)) N3 ((TI study OR AB study) OR (TI studies OR AB studies) OR (TI trial* OR AB trial*))) OR (((TI multicent* OR AB multicent*) OR (TI “multi-cent*” OR AB “multi-cent*”)) N3 ((TI study OR AB study) OR (TI studies OR AB studies) OR (TI trial* OR AB trial*))) OR (TI allocated OR AB allocated) OR (((TI “open label” OR AB “open label”) OR (TI “open-label” OR AB “open-label”)) N5 ((TI study OR AB study) OR (TI studies OR AB studies) OR (TI trial* OR AB trial*))) OR (((TI equivalence OR AB equivalence) OR (TI superiority OR AB superiority) OR (TI “non-inferiority” OR AB “non-inferiority”) OR (TI noninferiority OR AB noninferiority)) N3 ((TI study OR AB study) OR (TI studies OR AB studies) OR (TI trial* OR AB trial*))) OR ((TI “pragmatic study” OR AB “pragmatic study”) OR (TI “pragmatic studies” OR AB “pragmatic studies”)) OR (((TI pragmatic OR AB pragmatic) OR (TI practical OR AB practical)) N3 (TI trial* OR AB trial*)) OR (((TI quasiexperimental OR AB quasiexperimental) OR (TI “quasi-experimental” OR AB “quasi-experimental”)) N3 ((TI study OR AB study) OR (TI studies OR AB studies) OR (TI trial* OR AB trial*))) OR (TI trial)))

NOT

(((TI genetic* OR AB genetic*) OR (TI tau OR AB tau) OR (TI amyloid* OR AB amyloid*) OR (TI SURGICAL* OR AB SURGICAL*) OR (TI SURGERY* OR AB SURGERY*) OR (TI PROTEOMIC* OR AB PROTEOMIC*) OR (TI GENOMIC* OR AB GENOMIC*) OR (TI PROTEIN* OR AB PROTEIN*) OR (TI biomarker* OR AB biomarker*) OR (TI Presenilin* OR AB Presenilin*) OR (TI Progranulins OR AB Progranulins) OR (TI peptid* OR AB peptid*) OR (TI Niemann-Pick OR AB Niemann-Pick)))

**CENTRAL/Cochrane Library**

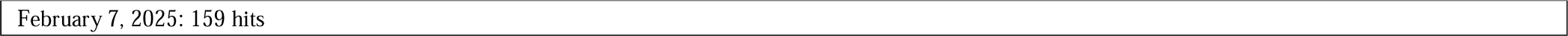

(((“young onset”:ti,ab OR “younger onset”:ti,ab OR “early onset”:ti,ab OR (young* NEXT “people”):ti,ab OR (young* NEXT person*):ti,ab OR frontotemporal:ti,ab) NEAR/3 (DEMENT*:ti,ab OR ALZHEIMER*:ti,ab)) OR ([mh “Frontotemporal Dementia”] OR FTD:ti,ab OR (pick* NEXT “disease”):ti,ab OR (Wilhelmsen* NEXT “Disease”):ti,ab))

NOT

(genetic*:ti,ab OR tau:ti,ab OR amyloid*:ti,ab OR SURGICAL*:ti,ab OR SURGERY*:ti,ab OR PROTEOMIC*:ti,ab OR GENOMIC*:ti,ab OR PROTEIN*:ti,ab OR biomarker*:ti,ab OR Presenilin*:ti,ab OR Progranulins:ti,ab OR peptid*:ti,ab OR Niemann-Pick:ti,ab)

NOT

((“clinicaltrials” OR “who.int” OR “www.clinicaltrialsregister.eu”):so OR conference proceeding:pt)

## Appendix 2. Excluded studies with exclusion reasons (full text stage; n=80; alphabetical order)

Aleixo M.A.R.; Santos R.L.; Dourado M.C.N. (2017): Efficacy of music therapy in the neuropsychiatric symptoms of dementia: Systematic review. In: *J. Bras. Psiquiatr.* 66 (1), 52 EP - 61-SP - 52 EP - 61. DOI: 10.1590/0047-2085000000150. Ineligible population (database search)

Aplaon M, Belchior P, Li L, et al. INTERVENTIONS FOR INDIVIDUALS WITH YOUNG-ONSET DEMENTIA. A REVIEW OF THE LITERATURE 2016: 1–4. Ineligible design (review search following citation search)

Aplaon M, Belchior P, Li L, et al. INTERVENTIONS FOR INDIVIDUALS WITH YOUNG-ONSET DEMENTIA. A REVIEW OF THE LITERATURE 2016: 1–4. Ineligible design (review search following citation search)

Appelhof B, Bakker C, Vugt ME de, et al. Effects of a Multidisciplinary Intervention on the Presence of Neuropsychiatric Symptoms and Psychotropic Drug Use in Nursing Home Residents WithYoung-Onset Dementia: Behavior and Evolution of Young-Onset Dementia Part 2 (BEYOND-II) Study. *Am J Geriatr Psychiatry* 2019; 27: 581–589. Duplicate / Report that was finally included (database search)

Appelhof B, Bakker C, Vugt ME de, et al. Effects of a Multidisciplinary Intervention on the Presence of Neuropsychiatric Symptoms and Psychotropic Drug Use in Nursing Home Residents WithYoung-Onset Dementia: Behavior and Evolution of Young-Onset Dementia Part 2 (BEYOND-II) Study. *Am J Geriatr Psychiatry* 2019; 27: 581–589. Duplicate (other sources)

Assogna M, Sprugnoli G, Press DZ, et al. Gamma-induction in frontotemporal dementia (GIFTeD) randomized placebo-controlled trial: Rationale, noninvasive brain stimulation protocol, and study design 2021; 7: NA-NA. Ineligible study design (citation search)

Bielderman A, van Corven CTM, Koopmans, Raymond T. C. M., et al. Evaluation of the SPAN intervention for people living with young-onset dementia in the community and their family caregivers: a randomized controlled trial. *Aging Ment Health* 2024; 28: 275–284. Duplicate / Report that was finally included (other sources)

Boots L, Vugt ME de, Kempen, Gertrudis I. J. M., et al. Effectiveness of the blended care self-management program “Partner in Balance” for early-stage dementia caregivers: study protocol for a randomized controlled trial 2016; 17: NA-NA. Ineligible population (citation search)

Boots L, Vugt ME de, Kempen, Gertrudis I. J. M., et al. Effectiveness of the blended care self-management program “Partner in Balance” for early-stage dementia caregivers: study protocol for a randomized controlled trial 2016; 17: NA-NA. Duplicate (citation search)

Boots L, Vugt ME de, Kempen, Gertrudis I. J. M., et al. Effectiveness of the blended care self-management program “Partner in Balance” for early-stage dementia caregivers: study protocol for a randomized controlled trial 2016; 17: NA-NA. Duplicate (citation search)

Boots L, Vugt ME de, Smeets CHW, et al. Implementation of the Blended Care Self-Management Program for Caregivers of People With Early-Stage Dementia (Partner in Balance): Process Evaluation of a Randomized Controlled Trial 2017; 19: e423-e423. Ineligible population (citation search)

Bruinsma J. Family caregivers of persons with young-onset dementia 2022: NA-NA. Ineligible study design (citation search)

Bruinsma J, Peetoom K, Boots L, et al. Tailoring the web-based ‘Partner in Balance’ intervention to support spouses of persons with frontotemporal dementia 2021; 26: 100442. Ineligible study design (citation search)

Bruinsma J, Peetoom K, Boots L, et al. Tailoring the web-based ‘Partner in Balance’ intervention to support spouses of persons with frontotemporal dementia 2021; 26: 100442. Duplicate (citation search)

Cations M, Withall A, Horsfall R, et al. Why aren’t people with young onset dementia and their supporters using formal services? Results from the INSPIRED study 2017; 12: e0180935-e0180935. Ineligible study design (citation search)

Cations M, Withall A, Horsfall R, et al. Why aren’t people with young onset dementia and their supporters using formal services? Results from the INSPIRED study 2017; 12: e0180935-e0180935. Duplicate (citation search)

Clare, Linda; Teale, Julia C.; Toms, Gill; Kudlicka, Aleksandra; Evans, Isobel; Abrahams, Sharon et al. (2018): Cognitive rehabilitation, self-management, psychotherapeutic and caregiver support interventions in progressive neurodegenerative conditions: A scoping review. In: *NeuroRehabilitation* 43 (4), p. 443–471. DOI: 10.3233/NRE-172353. Ineligible population (database search)

Climans, Renee; Berall, Anna; Santiago, Anna Theresa; Gardner, Sandra; Margles, Donna; Shnall, Adriana (2023): Evaluation of virtual caregiver support groups for young-onset dementia family caregivers. In: *Social Work with Groups* 46 (2), p. 157–170. DOI: 10.1080/01609513.2022.2148038. Ineligible study design (database search)

Climans R, Berall A, Santiago AT, et al. Evaluation of virtual caregiver support groups for young-onset dementia family caregivers. *Social Work with Groups* 2023; 46: 157–170. Ineligible study design (other sources)

Craig D and Strivens E. Facing the times: A young onset dementia support group: Facebook^TM^ style 2016; 35: 48–53. Ineligible study design (citation search)

Daemen M, Bruinsma J, Bakker C et al. (2022) A cross-sectional evaluation of the Dutch RHAPSODY program: online information and support for caregivers of persons with young-onset dementia. Internet Interv 28:100530. DOI:10.1016/j.invent.2022.100530 Duplicate of a report that was finally included (citation search)

Diez J.M.B.; Subirana S.R.; Adell M.A.M. (2009): Evidences of the usefulness of a cognitive training program. In: FMC Formacion Med. Continuada Aten. Prim. 16 (7), 418 EP - 423-SP - 418 EP - 423. DOI: 10.1016/S1134-2072(09)71962-0. Foreign language (database search)

Dimitriou T.; Papatriantafyllou J.; Konsta A.; Kazis D.; Athanasiadis L.; Ioannidis P. et al. (2022): Assess of Combinations of Non-Pharmacological Interventions for the Reduction of Irritability in Patients with Dementia and their Caregivers: A Cross-Over RCT. In: *Brain Sci* 12 (6), p. 691. DOI: 10.3390/brainsci12060691. Ineligible population (database search)

Draper B, Cations M, White FA, et al. Time to diagnosis in young-onset dementia and its determinants: the INSPIRED study 2016; 31: 1217– 1224. Ineligible study design (citation search)

Dowling GA, Merrilees J, Mastick J, et al. Life enhancing activities for family caregivers of people with frontotemporal dementia. *Alzheimer Dis Assoc Disord* 2014; 28: 175–181. Duplicate / Report that was finally included (other sources)

Fisher A, Cheung SC, O’Connor C, et al. The Acceptability and Usefulness of Positive Behaviour Support Education for Family Carers of People With Frontotemporal Dementia: A Pilot Study 2022; 36: 73–83. Ineligible study design (citation search)

Fisher A, Cheung SC, O’Connor C, et al. The Acceptability and Usefulness of Positive Behaviour Support Education for Family Carers of People With Frontotemporal Dementia: A Pilot Study 2022; 36: 73–83. Duplicate (citation search)

Goldberg EL. Filling an unmet need: a support group for early stage/ young onset Alzheimer’s disease and related dementias 2011; 107: NA-NA. Ineligible study design (citation search)

Gossink, Flora; Pijnenburg, Yolande; Scheltens, Philip; Pera, Aafke; Kleverwal, Rikie; Korten, Nicole et al. (2018): An intervention programme for caregivers of dementia patients with frontal behavioural changes: an explorative study with controlled effect on sense of competence. In: *Psychogeriatrics* 18 (6), p. 451–459. DOI: 10.1111/psyg.12351. Ineligible population (database search)

Gu J.; Long W.; Zeng S.; Li C.; Fang C.; Zhang X. (2024): Neurologic music therapy for non-fluent aphasia: a systematic review and meta-analysis of randomized controlled trials. In: *Front Neurol* 15, p. 1395312. DOI: 10.3389/fneur.2024.1395312. Ineligible population (database search)

Greenwood N, Pelone F and Hassenkamp A-M. General practice based psychosocial interventions for supporting carers of people with dementia or stroke: a systematic review. *BMC Fam Pract* 2016; 17: 3. Ineligible population (other sources)

Grote V, Leong CA, Summers A, et al. Randomized controlled trial of a positive emotion regulation intervention to reduce stress in family caregivers of individuals with Alzheimer’s disease: Protocol and design for the LEAF 2.0 study 2023: NA-NA. Ineligible population (citation search)

Hewitt P, Watts CM, Hussey JS, et al. Does a Structured Gardening Programme Improve Well-Being in Young-Onset Dementia? A Preliminary Study 2013; 76: 355–361. Ineligible study design (citation search)

Hewitt P, Watts CM, Hussey JS, et al. Does a Structured Gardening Programme Improve Well-Being in Young-Onset Dementia? A Preliminary Study 2013; 76: 355–361. Duplicate (citation search)

Hooghiemstra AM, Eggermont LHP, Scheltens P, et al. Study protocol: EXERcise and cognition in sedentary adults with early-ONset dementia (EXERCISE-ON). *BMC Neurol* 2012; 12: 75. Duplicate (other sources)

Hooghiemstra, Astrid M.; Eggermont, Laura H. P.; Scheltens, Philip; van der Flier, Wiesje M.; Bakker, Jet; Greef, Mathieu H. G. de et al. (2012): Study protocol: EXERcise and cognition in sedentary adults with early-ONset dementia (EXERCISE-ON). In: *BMC Neurol* 12, p. 75. DOI: 10.1186/1471-2377-12-75. Duplicate / Seed reference for citation searching but has been later excluded because no article with findings could have been identified (other sources)

Kimura, Takemi; Takamatsu, Junichi; Fujii, M., Hatakeyama, R., Fukuoka, Y., et al., Fujii, Hirono, N., Mori, E., Ikejiri, Y., et al., Hirono (2013): Pilot study of pharmacological treatment for frontotemporal lobar degeneration: Effect of lavender aroma therapy on behavioral and psychological symptoms. In: *Geriatr Gerontol Int* 13 (2), p. 516–517. DOI: 10.1111/ggi.12025. Ineligible study design (database search)

Kinney JM, Kart CS and Reddecliff L. ‘That’s me, the Goother’: Evaluation of a program for individuals with early-onset dementia 2011; 10: 361–377. Ineligible study design (citation search)

Kortte KB and Rogalskı E. Behavioural interventions for enhancing life participation in behavioural variant frontotemporal dementia and primary progressive aphasia 2013; 25: 237–245. Ineligible design (review search following citation search)

Kurz A, Bakker C, Böhm M, Diehl-Schmid J, Dubois B, Ferreira C, Gage H, Graff C, Hergueta T, Jansen S, Jones B, Komar A, de Mendonça A, Metcalfe A, Milecka K, Millenaar J, Orrung Wallin A, Oyebode J, Schneider-Schelte H, Saxl S, de Vugt M. (2016): RHAPSODY - Internet-based support for caregivers of people with young onset dementia: program design and methods of a pilot study. In: *Int Psychogeriatr*, p. 1–9. DOI: 10.1017/S1041610216001186. Duplicate of a report that was finally included (database search)

Kurz A, Bakker C, Böhm M, et al. RHAPSODY - Internet-based support for caregivers of people with young onset dementia: program design and methods of a pilot study. *Int Psychogeriatr* 2016; 28: 2091–2099. Duplicate / Report that was finally included (other sources)

Loi, Samantha M.; Flynn, Libby; Cadwallader, Claire; Stretton-Smith, Phoebe; Bryant, Christina; Baker, Felicity A. (2022): Music and Psychology & Social Connections Program: Protocol for a Novel Intervention for Dyads Affected by Younger-Onset Dementia. In: *Brain Sci* 12 (4). DOI: 10.3390/brainsci12040503. Ineligible study design / Seed reference for citation searching but has been later excluded because no article with findings could have been identified (database search)

Metcalfe A, Jones B, Mayer J, et al. Online information and support for carers of people with young-onset dementia: A multi-site randomised controlled pilot study. *Int J Geriatr Psychiatry* 2019; 34: 1455–1464. Duplicate of a report that was finally included (other sources)

Marks, Genee; McVilly, Keith (2020): Trained assistance dogs for people with dementia: a systematic review. In: *Psychogeriatrics* 20 (4), p. 510–521. DOI: 10.1111/psyg.12529. Ineligible population (database search)

Mayrhofer A, Mathie E, McKeown J, et al. Age-appropriate services for people diagnosed with young onset dementia (YOD): a systematic review 2017; 22: 933–941. Ineligible design (review search following citation search)

Mayrhofer A, Mathie E, McKeown J, et al. Age-appropriate services for people diagnosed with young onset dementia (YOD): a systematic review 2017; 22: 933–941. Ineligible design (review search following citation search)

Mitchell ARK, Kelso W, Paynter C, et al. Peer Support for Caregivers of People Living with Posterior Cortical Atrophy in Melbourne, Australia: A Feasibility Study 2024; 21: 513. Ineligible design (citation search)

Mitchell ARK, Kelso W, Paynter C, et al. Peer Support for Caregivers of People Living with Posterior Cortical Atrophy in Melbourne, Australia: A Feasibility Study 2024; 21: 513. Duplicate (citation search)

Morgan D (2002): Review: pharmacological and non-pharmacological interventions improve outcomes in patients with dementia and their caregivers. In: *Evidence Based Nursing*, p. 20. Ineligible population (database search)

Moyle W, Spencer M, Qi M, et al. Telephone and online support programs and assistive technologies that support informal carers of people living with young-onset dementia: A systematic review 2025: NA-NA. Ineligible design (review search following citation search)

Moyle W, Spencer M, Qi M, et al. Telephone and online support programs and assistive technologies that support informal carers of people living with young-onset dementia: A systematic review 2025: NA-NA. Ineligible design (review search following citation search)

Mulders A, Fick IWF, Bor H, et al. Prevalence and Correlates of Neuropsychiatric Symptoms in Nursing Home Patients With Young-Onset Dementia: The BEYOnD Study 2016; 17: 495–500. Ineligible study design (citation search)

Mulders A, Zuidema SU, Verhey FRJ, et al. Characteristics of institutionalized young onset dementia patients – the BEYOnD study 2014; 26: 1973–1981. Ineligible study design (citation search)

Nakanishi K and Yamaga T. Effect of Instrumental Activities of Daily Living habituation due to routinising therapy in patients with frontotemporal dementia 2021; 14: e240167-e240167. Ineligible study design (citation search)

O’Connell ME, Crossley M, Cammer A, et al. Development and evaluation of a telehealth videoconferenced support group for rural spouses of individuals diagnosed with atypical early-onset dementias 2013; 13: 382–395. Ineligible study design (citation search)

O’Connell ME, Crossley M, Cammer A, et al. Development and evaluation of a telehealth videoconferenced support group for rural spouses of individuals diagnosed with atypical early-onset dementias 2013; 13: 382–395. Duplicate (UDCS)

O’Connell ME, Crossley M, Cammer A, et al. Development and evaluation of a telehealth videoconferenced support group for rural spouses of individuals diagnosed with atypical early-onset dementias. *Dementia* 2014; 13: 382–395. Ineligible study design (other sources)

O’Connor C, Mioshi E, Kaizik C, et al. Positive behaviour support in frontotemporal dementia: A pilot study 2020; 31: 507–530. Ineligible study design (citation search)

O’Connor C, Mioshi E, Kaizik C, et al. Positive behaviour support in frontotemporal dementia: A pilot study 2020; 31: 507–530. Duplicate (citation search)

O’Connor CM, Clemson L, Brodaty H, et al. The tailored activity program (TAP) to address behavioral disturbances in frontotemporal dementia: a feasibility and pilot study. *Disabil Rehabil* 2019; 41: 299–310. Duplicate / Report that was finally included (other sources)

Pac Soo V.; Baker F.A.; Sousa T.V.; Odell-Miller H.; Stensaeth K.; Wosch T. et al. (2023): Statistical analysis plan for HOMESIDE: a randomised controlled trial for home-based family caregiver-delivered music and reading interventions for people living with dementia. In: *Trials* 24 (1), p. 316. DOI: 10.1186/s13063-023-07327-8. Ineligible population (database search)

Park, Juyoung; Cohen, Iris (2019): Effects of exercise interventions in older adults with various types of dementia: Systematic review. In: *Activities, Adaptation & Aging* 43 (2), p. 83–117. DOI: 10.1080/01924788.2018.1493897. Ineligible population (database search)

Perin S, Lai R, Diehl-Schmid J, et al. Online counselling for family carers of people with young onset dementia: The RHAPSODY-Plus pilot study. *Digit Health* 2023; 9: 20552076231161962. Ineligible study design (other sources)

Perin S, Lai R, Diehl-Schmid J, et al. Online counselling for family carers of people with young onset dementia: The RHAPSODY-Plus pilot study 2023; 9: 205520762311619-205520762311619. Duplicate (citation search)

Perin S, Lai R, Diehl-Schmid J, et al. Online counselling for family carers of people with young onset dementia: The RHAPSODY-Plus pilot study 2023; 9: 205520762311619-205520762311619. Duplicate (citation search)

Rodriguez-Sanchez E, Patino-Alonso MC, Mora-Simón S, et al. Effects of a psychological intervention in a primary health care center for caregivers of dependent relatives: a randomized trial. *Gerontologist* 2013; 53: 397–406. Ineligible population (other sources)

Rogalski, Emily; Roberts, Angela; Salley, Elizabeth; Saxon, Marie; Fought, Angela; Esparza, Marissa et al. (2022): Communication Partner Engagement: A Relevant Factor for Functional Outcomes in Speech-Language Therapy for Aphasic Dementia. In: *J Gerontol B Psychol Sci Soc Sci* 77 (6), p. 1017–1025. DOI: 10.1093/geronb/gbab165. Ineligible population (database search)

Roheger, Mandy; Riemann, Steffen; Brauer, Andreas; McGowan, Ellen; Grittner, Ulrike; Floel, Agnes; Meinzer, Marcus (2024): Non-pharmacological interventions for improving language and communication in people with primary progressive aphasia. In: *Cochrane Database Syst Rev* 5, CD015067. DOI: 10.1002/14651858.CD015067.pub2. Ineligible population (database search)

Stefano M de, Esposito S, Iavarone A, et al. Effects of Phone-Based Psychological Intervention on Caregivers of Patients with Early-Onset Alzheimer’s Disease: A Six-Months Study during the COVID-19 Emergency in Italy. *Brain Sci* 2022; 12. Duplicate / Report that was finally included (other sources)

Shinagawa, Shunichiro; Nakajima, Shinichiro; Plitman, Eric; Graff-Guerrero, Ariel; Mimura, Masaru; Nakayama, Kazuhiko; Miller, Bruce L. (2015): Non-pharmacological management for patients with frontotemporal dementia: a systematic review. In: *J Alzheimers Dis* 45 (1), p. 283–293. DOI: 10.3233/JAD-142109. Ineligible study design (database search)

Shinagawa, Shunichiro; Nakajima, Shinichiro; Plitman, Eric; Graff-Guerrero, Ariel; Mimura, Masaru; Nakayama, Kazuhiko et al. (2015): Non-pharmacological management for patients with frontotemporal dementia: A systematic review. In: *J Alzheimers Dis* 45 (1), p. 283–293. DOI: 10.1016/j.archger.2012.04.011. Duplicate (database search)

Sullivan MP, Williams V, Grillo A, et al. Peer support for people living with rare or young onset dementia: An integrative review 2022; 21: 2700–2726. Ineligible design (review search following citation search)

Sullivan MP, Williams V, Grillo A, et al. Peer support for people living with rare or young onset dementia: An integrative review 2022; 21: 2700–2726. Ineligible design (review search following citation search)

van Berg E den, Papma JM, van der Tholen FC, et al. Mindfulness-Based Stress Reduction in Pre-symptomatic Genetic Frontotemporal Dementia: A Pilot Study 2022; 13: NA-NA. Ineligible study design (RICS)

Vlotinou P, Tsiakiri A, Detsaridou G, et al. Occupational Therapy Interventions in Patients with Frontotemporal Dementia: A Systematic Review 2023; 11: 71. Ineligible design (review search following citation search)

Vlotinou P, Tsiakiri A, Detsaridou G, et al. Occupational Therapy Interventions in Patients with Frontotemporal Dementia: A Systematic Review 2023; 11: 71. Ineligible design (review search following citation search)

Volkmer A.; Walton H.; Swinburn K.; Spector A.; Warren J.D.; Beeke S. (2023): Results from a randomised controlled pilot study of the Better Conversations with Primary Progressive Aphasia (BCPPA) communication partner training program for people with PPA and their communication partners. In: *Pilot feasibility stud.* 9 (1), p. 87. DOI: 10.1186/s40814-023-01301-6. Ineligible population (database search)

Waddington C, Harding E, Brotherhood E, et al. The Development of Videoconference-Based Support for People Living With Rare Dementias and Their Carers: Protocol for a 3-Phase Support Group Evaluation (Preprint) 2021: NA-NA. Ineligible study design (citation search)

Waddington C, Harding E, Brotherhood E, et al. The Development of Videoconference-Based Support for People Living With Rare Dementias and Their Carers: Protocol for a 3-Phase Support Group Evaluation 2022; 11: e35376-e35376. Ineligible study design (citation search)

Windle G.; Flynn G.; Hoare Z.; Goulden N.; Tudor Edwards R.; Anthony B. et al. (2025): Evaluating the effects of the World Health Organization’s online intervention ‘iSupport’ to reduce depression and distress in dementia carers: a multi-centre six-month randomised controlled trial in the UK. In: *Lancet. Reg. Health. Eur.* 48, p. 101125. DOI: 10.1016/j.lanepe.2024.101125. Ineligible population (database search)

## Appendix 3. Raw data set

See separate spreadsheet.

## Notes

### Competing Interest Statement

The authors have declared no competing interest.

